# Long-term Coexistence of Severe Acute Respiratory Syndrome Coronavirus 2 (SARS-CoV-2) with Antibody Response in Coronavirus Disease 2019 (COVID-19) Patients

**DOI:** 10.1101/2020.04.13.20040980

**Authors:** Bin Wang, Li Wang, Xianggen Kong, Jin Geng, Di Xiao, Chunhong Ma, Xue-Mei Jiang, Pei-Hui Wang

**Affiliations:** Jinan Infectious Diseases Hospital of Shandong University, Jinan, Shandong 250021, China; Advanced Medical Research Institute, Shandong University, Jinan, Shandong 250012, China

**Keywords:** SARS-CoV-2, COVID-19, adaptive immunity, innate immunity, IgG antibody

## Abstract

Severe acute respiratory syndrome coronavirus 2 infection causing coronavirus disease 2019 has spread worldwide. Whether antibodies are important for the adaptive immune responses against SARS-CoV-2 infection needs to be determined. Here, 26 cases of COVID-19 in Jinan, China, were examined and shown to be mild or with common clinical symptoms and no cases of severe symptoms were found among these patients. A striking feature of some patients is that SARS-CoV-2 could exist in patients who have virus-specific IgG antibodies for a very long period, with two cases for up to 50 days. One COVID-19 patient who did not produce any SARS-CoV-2-bound IgG successfully cleared SARS-CoV-2 after 46 days of illness, revealing that without antibody-mediated adaptive immunity, innate immunity may still be powerful enough to eliminate SARS-CoV-2. Overall, this report may provide a basis for further analysis of both innate and adaptive immunity in SARS-CoV-2 clearance, especially in non-severe cases. This study also has implications for understanding the pathogenesis and treatment of SARS-CoV-2.

---

The first severe acute respiratory syndrome coronavirus 2 (SARS-CoV-2) outbreak was reported in December 2019, and the virus has rapidly spread worldwide within 3 months (Wu et al., 2020; Zhou et al., 2020; Zhu et al., 2020). The importance of innate and adaptive immunity in the defense against SARS-CoV-1 needs to be urgently determined (Thevarajan et al., 2020). To fulfill the pressing need, we examined antibody generation and virus clearance in 26 patients with SARS-CoV-2-induced coronavirus disease 2019 (COVID-19) in Jinan, China.

A total of 26 patients from 5 to 72 years old were determined to be SARS-CoV-2 RNA positive by sputum, stool, or nasopharyngeal swabs. The clinical characteristics of the patients and chest CT scans were also examined. According to the “Fifth Revised Trial Version of the Novel Coronavirus Pneumonia Diagnosis and Treatment Guidance” (http://www.nhc.gov.cn/yzygj/s7652m/202002/41c3142b38b84ec4a748e60773cf9d4f.shtml), all of them are non-severe COVID-19 patients (**Table 1**).

**Table 1.** Clinical characteristics of the 26 hospitalized SARS-CoV-2 patients and corresponding timelines of IgG antibody production.

| Patients | Gender/Age (Y.) | Type | IgG P. (D.) | LDVP/specimens | Co-existence (D.) |
| --- | --- | --- | --- | --- | --- |
| 1 | F/18-60 | C | 22 | 22/NP | 0 |
| 2 | M/18-60 | C | 7 | 57/St | 50 |
| 3 | F/18-60 | C | 23 | 18/NP | NA |
| 4 | F/18-60 | C | 16 | 32/St | 16 |
| 5 | F/18-60 | C | 23 | 29/NP | 6 |
| 6 | F/18-60 | C | 21 | 21/St | 0 |
| 7 | F/18-60 | M | 9 | 8/NP | NA |
| 8 | M/18-60 | C | 23 | 73/NP | 50 |
| 9 | M/18-60 | C | 10 | 23/NP | 13 |
| 10 | M/>60 | C | 9 | 19/Sp&NP | 10 |
| 11 | M/18-60 | C | 17 | 20/NP | 3 |
| 12 | F/<18 | M | 14 | 28/St | 14 |
| 13 | M/18-60 | C | 15 | 51/St | 36 |
| 14 | F/18-60 | C | 10 | 25/SP | 24 |
| 15 | M/18-60 | C | 24 | 36/NP | 12 |
| 16 | F/18-60 | C | 15 | 60/NP | 45 |
| 17 | M/18-60 | C | 20 | 36/SP | 16 |
| 18 | M/18-60 | M | 17 | 24/NP | 7 |
| 19 | M/18-60 | C | 12 | 21/SP | 9 |
| 20 | F/18-60 | C | 18 | 20/NP | 2 |
| 21 | F/>60 | C | 14 | 20/NP | 6 |
| 22 | M/18-60 | M | 10 | 7/NP | NA |
| 23 | F/18-60 | C | 15 | 18/NP | 3 |
| 24 | F/18-60 | C | 18 | 17/NP | NA |
| 25 | F/<18 | C | 10 | 23/St | 13 |
| 26 | F/<18 | M | ND* ( 66<sup>th</sup>) | 46/St | NA |

**Y:** year; **IgG P:** IgG positive; **D:** Day; **LDP:** Last day post-illness when SARS-CoV-2 nucleic acid testing shows positive; **NP:** nasopharyngeal; **Sp:** sputum; **St:** stool. The severity of COVID-19 was judged according to the “Fifth Revised Trial Version of the Novel Coronavirus Pneumonia Diagnosis and Treatment Guidance” (http://www.nhc.gov.cn/yzygj/s7652m/202002/41c3142b38b84ec4a748e60773cf9d4f.shtml). **M:** mild type, the clinical symptoms were mild and no pneumonia was found in imaging. **C:** common type, with fever, respiratory tract and other symptoms, the manifestations of pneumonia can be seen on imaging. **NA:** not applicable; **ND:** not detectable. \* IgG antibodies keep undetectable till 66th day post-illness for patient 26.

Immunoglobulin G (IgG) antibodies act as an indicator of recent and past infection, while IgM antibodies indicate current infection. SARS-CoV-2-bound IgG antibodies were detected in the serum of patients. For patients 3, 7, 22 and 24 (**Table 1**), SARS-CoV-2 nucleic acid measurements became negative before antibodies were detected. Although IgG antibody testing was positive for the first trial in all 4 patients, we could not determine the exact role of IgG antibodies in SARS-CoV-2 clearance because innate immunity is also sufficient to eliminate this virus (refer to the case of patient 26). However, the results did reveal that the antibodies are produced early and SARS-CoV-2 is cleared up, indicating that the immediate production of antibodies may contribute to virus clearance.

Interestingly, we observed that specimens from patients 2, 8, 13, and 16 who had been confirmed to be IgG positive still tested positive for SARS-CoV-2 nucleic acid for more than 35 days (**Table 1**), indicating that SARS-CoV-2 can coexist with its specific antibodies in the human body for an unexpectedly long time (36-50 days). According to the data collected from patient 2, IgG antibodies can be produced at least as early as the 7th day post illness. (**Table 1**). The average number of days for SARS-CoV-2-bound IgG antibodies to be first detected in the 4 patients was 15; thus, the early production of antibodies does not mean early elimination of this virus. The production of specific antibodies is believed to be effective for virus clearance (Jiang, 2020; Lu, 2020), but we could not reach such a conclusion from these cases. We did not observe a correlation between early adaptive immune responses and better clinical outcomes. Perhaps the specificity and titer of antibodies are more important. To our knowledge, to date, this is the longest period (36-50 days) to observe the coexistence of SARS-CoV-2 with its specific antibodies in COVID-19 patients. How this virus can circulate in the presence of specific antibodies for such a long time is an interesting question. Whether SARS-CoV-2 can act as HCV that have developed strategies to subvert humoral immunity and persists in the body is worth further investigation (Elsner et al., 2015; Fafi-Kremer et al., 2012; Takaya et al., 2019). These 3 patients were all 23-39−year-old adults, which may suggest that young adult individuals do not have obvious advantages in the early production of antibodies and the clearance of SARS-CoV-2 compared to older persons (patients 10 and 21).

We observed that patient 26, a 5-10−year-old female, was SARS-CoV-2 nucleic acid positive in a stool sample after 46 days of illness but became nucleic acid testing negative in specimens of sputum, stool, and nasopharyngeal swabs on day 47 post illness (**Table 1**). No SARS-CoV-2−specific antibodies were detected in the patient’s serum until the last sample collection day, which was the 66th day post illness. Although we did not collect data about virus-specific cellular immunity, it is known that cellular immunity is generated concomitantly with humoral immunity, so we could preliminarily exclude the potential role of cellular immunity in SARS-CoV-2 elimination in this case. Thus, from the data on patient 26, we may conclude that innate immunity, the first line of host defense, can play an essential role in SARS-CoV-2 clearance; moreover, innate immunity alone might be enough to clear the virus. This case may also indicate that some individuals may not generate specific antibodies after infection with SARS-CoV-2; thus, only testing SARS-CoV-2−specific antibodies is not a good standard to determine infection, but combination with the nucleic acid testing method may improve the accuracy of SARS-CoV-2 detection.

Patient 25, another 5-10 year-old female, was found to be IgG antibody positive on the 10th day post illness, and the patient turned SARS-CoV-2 nucleic acid negative on 23rd day post illness (**Table 1**). We also observed that a 5-10 year-old female patient (patient 7) produced IgG antibodies on the 14th day post illness, and this patient turned SARS-CoV-2 nucleic acid negative on the 28th day post illness (**Table 1**). These 2 cases may reveal that children do not show any defects in antibody production and SARS-CoV-2 elimination compared with adults.

Although a vaccine is believed to be the ultimate preventive measure against SARS-CoV-2 spread, generating a broadly protective and universal vaccine can take a long time (Lu, 2020). In this study, in the case of patient 26 (**Table 1**), we observed that innate immunity alone may be enough to completely clear SARS-CoV-2 infection. This is the first report that innate immunity plays such an essential role in the host defense against SARS-CoV-2, which highlights the importance of innate immunity in SARS-CoV-2 clearance. Further studies are required to determine which factors or signaling pathways of innate immunity contribute to this process. Whether individuals with such responses are still at risk for reinfection needs further exploration. Similar to other RNA viruses, SARS-CoV-2 RNA should be detected by endosomal Toll-like receptors (TLRs) and cytosolic RIG-I-like receptors or other RNA sensors that activate NLRP3 signaling, leading to the production of IFNs, ISGs, and proinflammatory cytokines (Kasumba and Grandvaux, 2019). Boosting innate immune signaling pathways, such as the TLR3 and RIG-I pathways, by drugs that mimic viral RNA may contribute to SARS-CoV-2 clearance (Kasumba and Grandvaux, 2019). However, these immune stimulators also induce an inflammatory response that may be harmful to patients; thus, antiviral immunity should be maximized, and inflammatory responses must be minimized. The inflammatory response that is responsible for the accumulation of cells and fluids contributes to SARS□induced lung injury (Huang et al., 2020; Wang et al., 2020). As immune stimulators, viral RNA mimics also induce an inflammatory response that may be harmful to patients; thus, a strategy, in which antiviral immunity should be maximized and inflammatory responses must be minimized, is challenging. Vaccine combined with innate immune stimulators may be more effective for fast SARS-CoV-2 clearance. We propose that the importance of innate immunity should be investigated further and that the titer and specificity of SARS-CoV-2−specific antibodies are important and should be seriously considered in vaccine development.

Our study is limited by the current reagents used in this study, which cannot be used to determine the titer and specificity of human antibodies against SARS-CoV-2. The titer of specific antibodies correlated with clinical outcomes remains to be investigated. The long-term coexistence of IgG antibody with SARS-CoV-2 in the human body raises the question of whether patients with antibodies are still at risk for reinfection. Our follow-up studies may answer this question and would, therefore, be beneficial to vaccine development. Second, we did not collect the earliest serum of patients, and we could not determine on which day the SARS-CoV-2 bound IgG antibodies were generated. Thus, we could not observe the dynamic changes in IgG antibodies during illness and recovery. Third, we lack severe patients as controls; therefore, we do not know whether the adaptive immunity of severe patients can respond earlier because of their strong immune response and whether they can clear the virus faster after antibodies are produced. Despite that, our study provided several novel pieces of information about the innate and adaptive immune response against SARS-CoV-2: SARS-CoV-2 bound IgG antibodies can be generated as early as 7 days post illness but can coexist with SARS-CoV-2 in patients long-term (up to 50 days); without SARS-CoV-2 bound IgG antibodies, innate immunity can also successfully clear this virus.

## Materials and Methods

Specimens from sputum, stool, and nasopharyngeal swabs were collected throughout the illness from January 30, 2020, to April 5, 2020. Viral RNA was extracted from clinical specimens and real-time reverse transcription-PCR (rRT-PCR) was performed to test the presence of SARS-CoV-2 using “Novel coronavirus 2019-nCoV nucleic acid detection kit” (Shanghai BioGerm Medical Biotechnology Co.,Ltd, China). The serum was collected at distinctive time points, and SARS-CoV-2−specific antibodies were detected using “New Coronavirus (2019-nCoV) Antibody Detection Kit” (INNOVITA, China). This study was approved by the ethics commissions of Jinan infectious disease hospital, Shandong, China.

## Data Availability

All data referred to in the manuscript are available.

## Acknowledgement

This work was supported by grants from COVID-19 emergency tackling research project of Shandong University (Grant No. 2020XGB03 to P.-H.W).

